# Antiviral treatments lead to the rapid accrual of hundreds of SARS-CoV-2 mutations in immunocompromised patients

**DOI:** 10.1101/2022.12.21.22283811

**Authors:** Nicholas M. Fountain-Jones, Robert Vanhaeften, Jan Williamson, Janelle Maskell, I-Ly J Chua, Michael Charleston, Louise Cooley

## Abstract

The antiviral Molnupiravir (Lageviro) is widely used across the world to treat SARS-CoV-2 infection. Molnupiravir reduces viral replication by inducing mutations throughout the genome, yet in patients that do not clear the infection, the longer-term impact of the drug on virus evolution is unclear. Here, we used a case-control approach to monitor SARS-CoV-2 genomes through time in nine immunocompromised -patients with five treated with Molnupiravir. Within days of treatment, we detected a large number of low-frequency mutations in patients and that these new mutations could persist and, in some cases, were fixed in the virus population. All patients treated with the drug accrued new mutations in the spike protein of the virus, including non-synonymous mutations that altered the amino acid sequence. Our study demonstrates that this commonly used antiviral can ‘supercharge’ viral evolution in immunocompromised patients, potentially generating new variants and prolonging the pandemic.

## Main text

Persistent SARS-CoV-2 infection in immunocompromised patients is recognized as an important source of genomic variation and is linked to the evolution of novel variants ^1–3^. How commonly used antivirals such as Molnupiravir shape viral evolution is an important knowledge gap. As of December 2022, Molnupiravir is routinely used globally to treat COVID patients in and outside of hospital settings, including treatment of immunocompromised patients ^4^. Molnupiravir and other similar direct-acting SARS-CoV-2 antivirals promote mutagenesis by incorporating the prodrug NHC (β-D-N4-hydroxycytidine) into the virus-dependent RNA polymerase (RdRp)^5^. When the RdRp uses the NHC modified RNA as a template it promotes an ‘error catastrophe’ ^5,6^ that inhibits the replication of SARS-CoV-2. For immunocompromised patients that do not clear infection after antiviral treatment, the impact that these drugs have on patterns of virus evolution is unclear.

We intensively monitored nine immunocompromised patients; five patients were swabbed pre and post Molnupiravir treatment and four patients did not receive the drug. We found that as little as 10 days after Molnupiravir treatment, patient SARS-CoV-2 genomes had accrued on average 30 new low-mid frequency variants (between 10-90% of reads, Fig 1). We did not find similar changes to viral diversity in the patients not treated with Molnupiravir (Fig. 1).

**Fig. 1:**
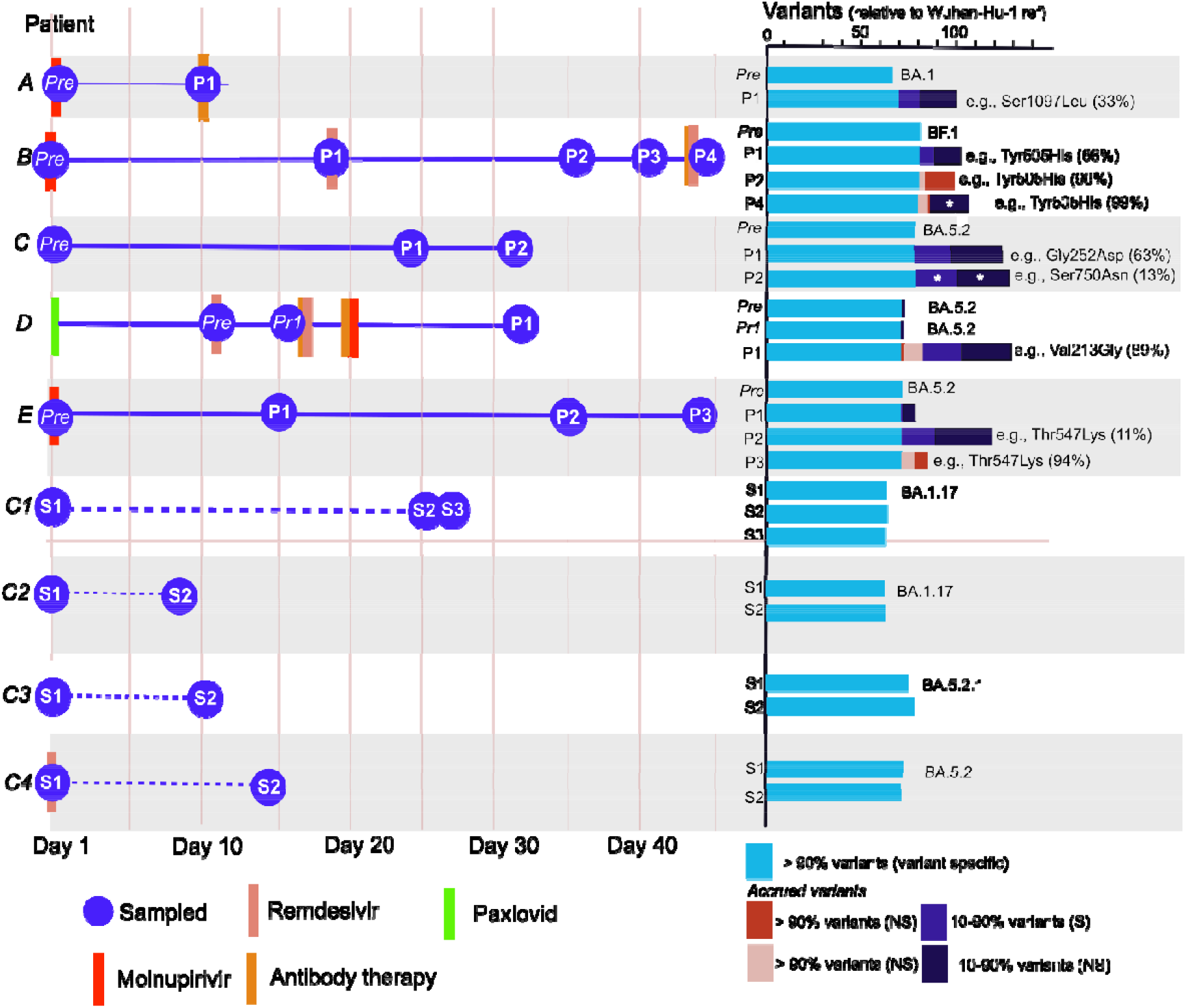
Patient histories, sampling regimen and variant count in each sample (Prep: pre-treatment sample, pr1: pre Molnupirivir treatment but post Remdesivir treatment, P1-4: subsequent samples post-treatment, S1-3: samples from untreated patients). The Omicron subvariant lineage of each patient is reported next to the variant count from the pre-sample. Patients with dotted lines (C1-4) were not treated with Molnupirivir. The text next to the variant counts highlights a non-synonymous mutation to the spike protein that had either the highest frequency or increased in frequency in subsequent samples. White star: variants found were completely distinct from the previous sample.

We commonly detected non-synonymous mutations in the spike protein that is the current vaccine target (Fig. 1, Appendix 1) and subsequent samples from these patients demonstrated that some of these spike mutations were fixed (frequency > 90%, e.g., Patient E Thr547Lys). In Patient B, 36 days after treatment 10 novel non-synonymous mutations were fixed, including mutations in the spike protein (Fig. 1/S1). This represents approximately 33% of the mutations that define the Omicron variant ^7^. However, we found some mutations were transient, as many were not detected in the subsequent samples (Fig. 1, multiple infections were ruled out).

These data highlight the risk of treating immunocompromised patients with error generating antivirals such as Molnupiravir. All of the individuals in our study remained persistently PCR positive post-treatment, although active monitoring for clearance was not undertaken by the institution. It is possible they were infectious in the hospital and in their communities, and onward transmission of these highly divergent viruses is likely. This commonly used class of antivirals has the capability to supercharge SARS-CoV-2 evolution, and uncontrolled use may generate new variants with a transmission advantage that prolongs the pandemic and makes other therapeutics less effective.

## Data Availability

All data will be available online at GISAID. The sequence alignment is available upon request.

## Data availability statement

All data will be available online at GISAID. The sequence alignment is available upon request.

## Supplementary text

### Methods

#### Study population

The 9 patients included in this study were immunocompromised due to a number of aetiologies, including haematological malignancy, solid organ and allogenic stem cell transplantation, and treatment of vasculitis that had tested qPCR positive to SARS CoV-2 infection. Six patients were treated with antivirals (one just with Remdesivir) and five were treated with Molnupirivir. The patients not treated with Molnupirivir were diagnosed with the virus before approval of the drug in Australia and acted as a quasi-control.

#### Real-Time PCR assay

Flocked swabs were collected from the throat and nasopharynx and immediately placed in 2.5mL viral transport Fluid (VTF).

Samples were vortexed for 1 minute and 200μL VTF was extracted on the Magnapure96 instrument (Roche) using the DNA/RNA SV kit and Pathogen Universal 200 protocol.

Ten microlitres of eluate was used in a 20μL multiplex real time PCR containing 1x MDX-016 mastermix (Meridian Bioscience), with primers and probes (Bioneer) targeting the SARS-CoV-2 matrix and RdRP genes, and human RNaseP gene as a control. Oligo sequences are shown below.

**Table S1.**
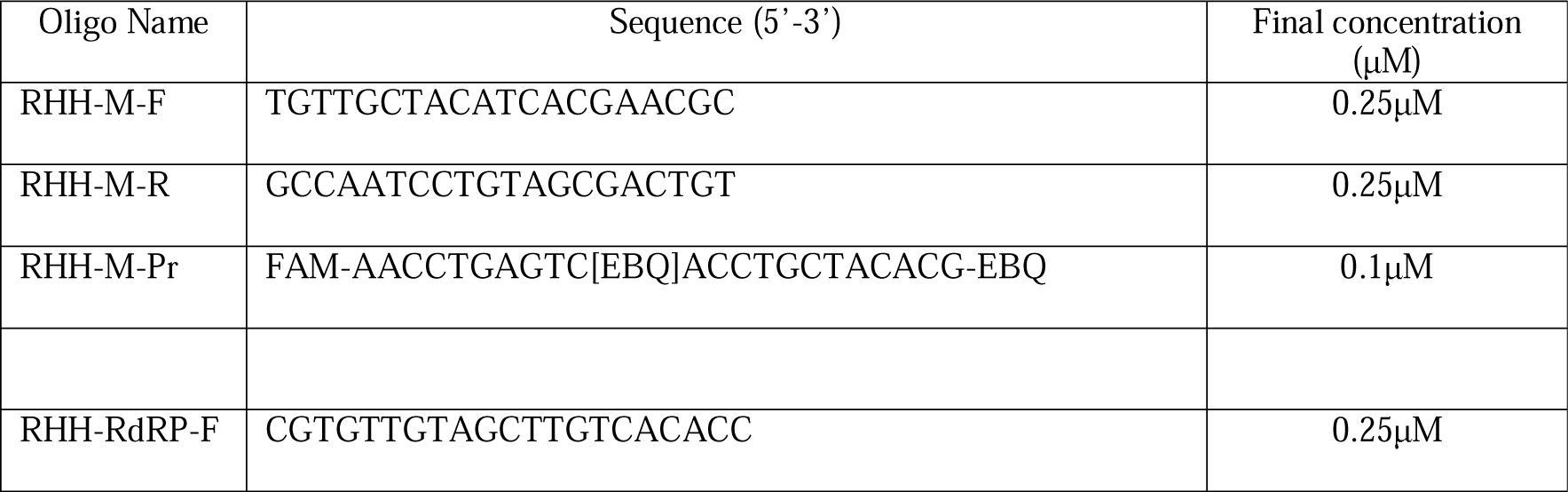
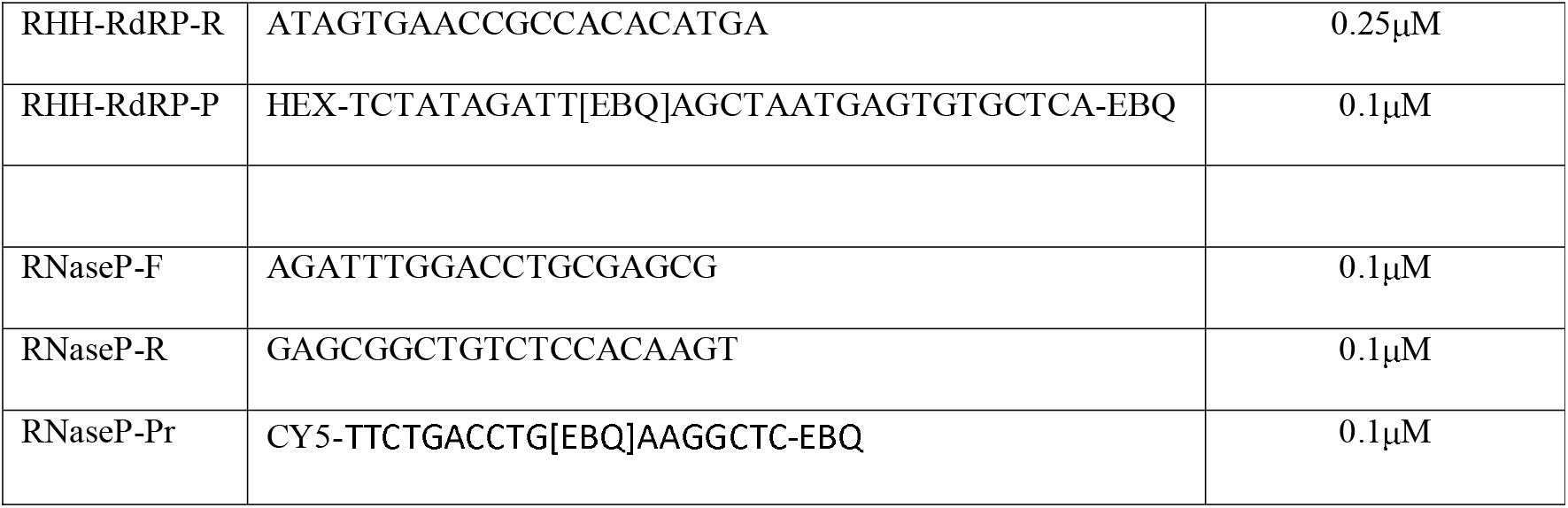
RT-PCR primer and probe sequences.

PCR was performed on an LC480 instrument (Roche) with thermal cycling parameters as follows: 50°C for 15 minutes, 95°C for 2 minutes, then 40 cycles of 95°C for 10 seconds and 60°C for 30 seconds with fluorescence collected at the 60°C step.

#### Tiled amplicon sequencing

Overlapping amplicons were generated using the QIAseq SARS-CoV-2 Primer Panel kit (Qiagen) as per the manufacturer’s instructions except substituting a custom primer set and using a 60°C annealing/extension temperature.

**Table S2.**
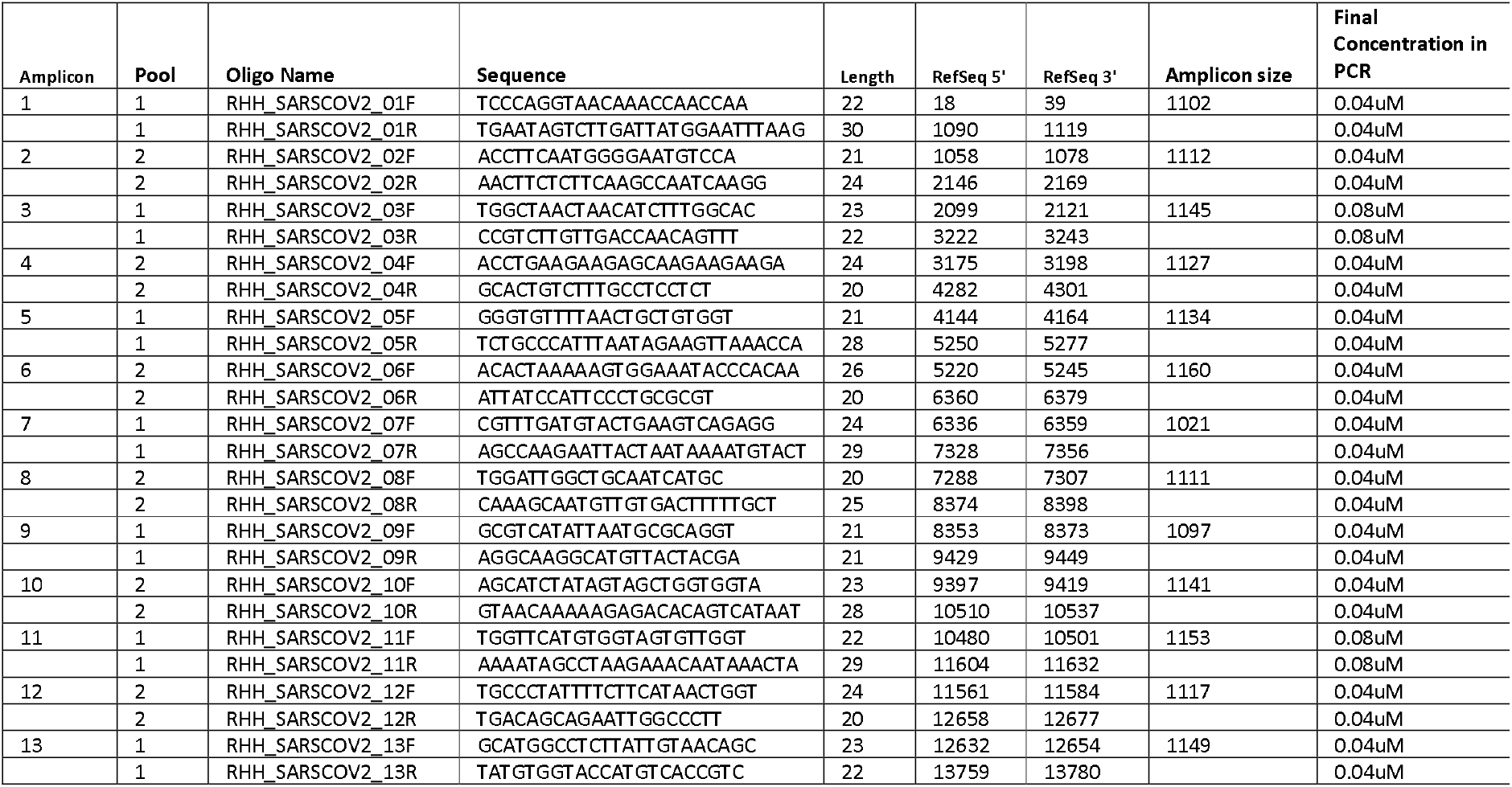
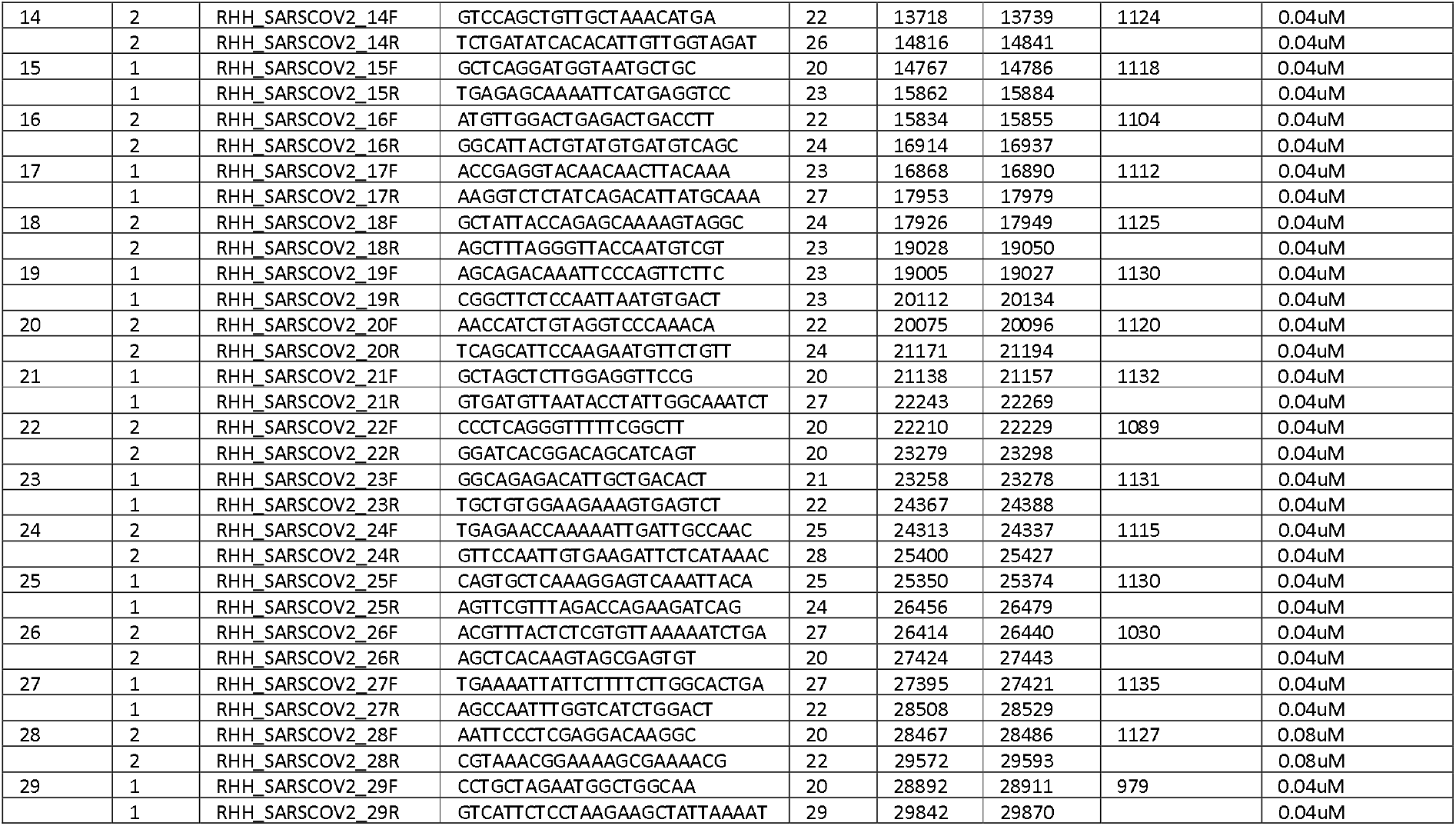
Tiled amplicon primer sequences.

Amplicon concentration was measured using the Qubit 1X BR kit on a Qubit flex instrument (Thermo). Approximately 200ng of amplicon was fragmented and barcoded using the Illumina DNA Prep kit as per the manufacturer’s instructions before 2×151 sequencing on an Illumina Miniseq. Basecalling, demultiplexing and adapter trimming were performed on-instrument.

#### Sequence analysis and variant calling

Raw reads were imported in CLC Genomics Workbench 21.0.5 and analyzed using a custom workflow. Reads were quality trimmed (<20) before being aligned to the SAR CoV-2 Wuhan-Hu-1 reference sequence. Primer sequences were trimmed from read mappings and variants called using the low frequency variant caller with a threshold of 10% VAF, a minimum coverage of 10-fold and a minimum of 5 variant read support. A consensus sequence was derived from mapped reads with bases below 10-fold coverage replaced with N and mixed base sites between 10% and 90% given IUB mixed base codes. Lineage and sub-lineages were identified through the web base Pangolin version 3.1.20 version date 28/02/2022.

To compare variants from our patient samples with global reference sequences and to visualize the locations of the mutations across the genome we used the Ultrafast Sample placement on Existing tRee (UShER) pipeline ^8^ and the University of California Santa Cruz (UCSC) genome viewer (https://genome.ucsc.edu/index.html).

#### Supplementary results

The variants that accrued in patients after treatment with Lageviro tended to be scattered across the genome and included mutations not commonly sampled in global Omicron genomes (Fig. S1). The accrued mutations we detected in the spike (s) gene tended to be clustered in two locations (Fig. S1), but the functional relevance of the mutations was unclear. While we did not detect any known drug resistance mutations, we did detect non-synonymous mutations in ORF 1b in the neighbouring amino acids (Fig. S1).

The UShER analysis showed that while most samples from individual patients clustered together there were potentially novel or rare mutations in the sequences post-treatment (Fig. 1). There were samples with so many mutations that they were phylogenetically distinct and difficult to place on the global SARS CoV-2 phylogeny. For example, the genomes from the 4^th^ sample from Patient B (P4) and the first sample from patient D (P1) were so distinct that the minimum number of mutations needed to add the sample to the global tree was 14 and 17 respectively (Fig. 1). While the first sample taken from Patient A (P1) could be added to the global reference tree without requiring any new mutations, these mutations accrued were phylogenetically distinct indicating that the sample was phylogenetically distinct from the pre-treatment sample prior and to the other samples taken post treatment (Fig. 1).

**Fig. S1:**
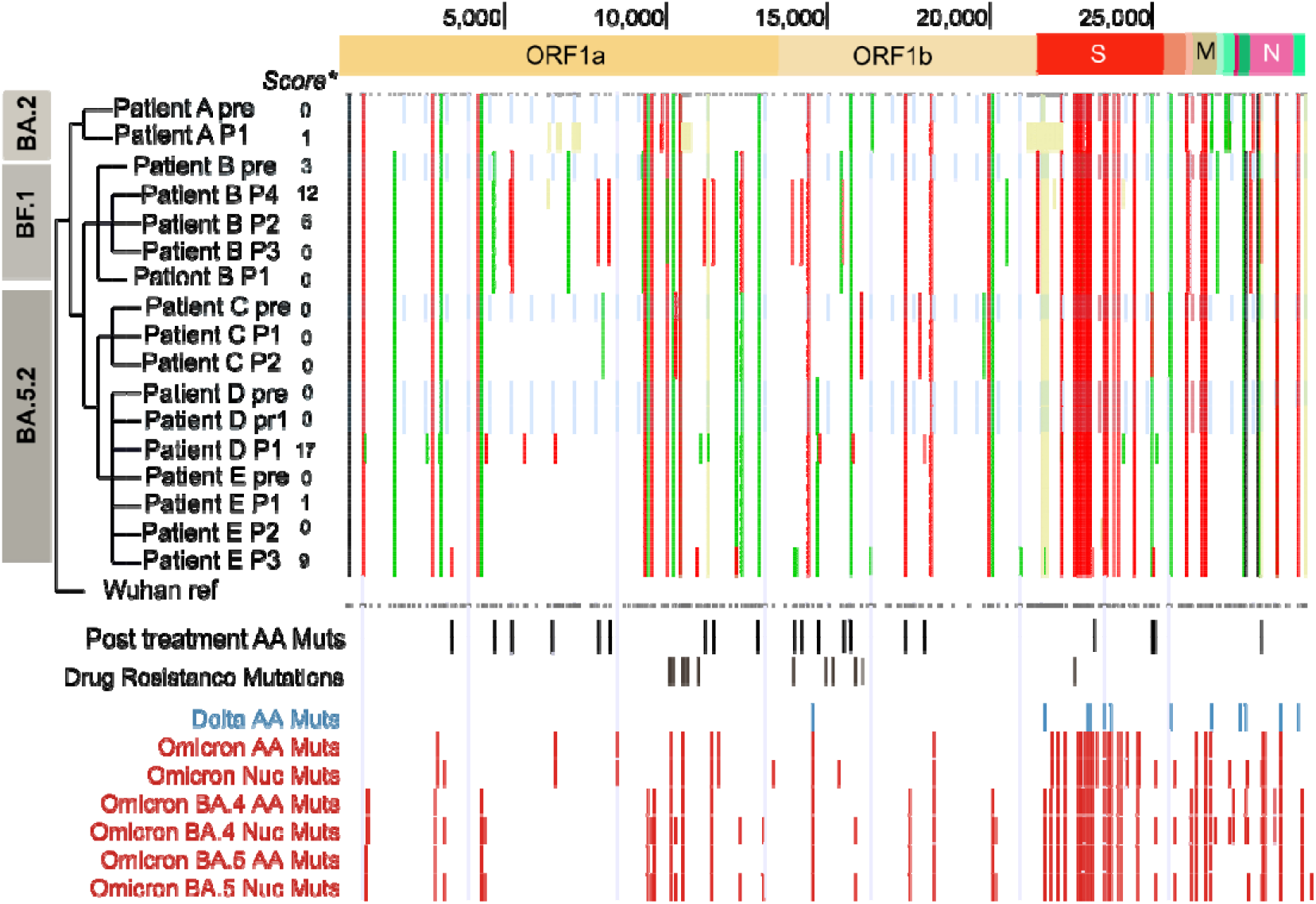
Waterfall plot showing the SARS CoV-2 genome coordinates of the nonsynonymous (amino acid (AA), red) and synonymous mutations (nucleotide (Nuc), green) across the treated patients in this study (top panel). Mid panels = Location of non-synonymous mutations just detected in treated patients with the location of known resistance mutations below. Bottom panels: Mutations defining omicron and BA.4/5. Light blue bars: Samples taken prior to Molnupirivir treatment. We used the Ultrafast Sample placement on Existing tRee (UShER) pipeline ^8^ to generate lineage assignments, phylogenetic placement and parsimony scores (left panel). The parsimony score is based on the minimum number of additional mutations needed to place the sequence in the reference tree. Wuhan ref = Wuhan reference sequence (MN908947.3).

